# Differences in US Adult Dietary Patterns by Cardiovascular Health and Socioeconomic Vulnerability

**DOI:** 10.1101/2025.01.02.25319924

**Authors:** Eric J. Brandt, Cindy Leung, Tammy Chang, John Z. Ayanian, Mousumi Banerjee, Matthias Kirch, Dariush Mozaffarian, Brahmajee K. Nallamothu

**Author notes:** Corresponding Author and Reprint Contact: Eric J Brandt, MD, MHS, FACC, 24 Frank Lloyd Wright Dr, Lobby A, Ann Arbor, MI 48106, USA; Fax: n/a. (cleung2hsph.harvard.edu). Note: Mozaffarian and Nallamothu are co-senior authors. **Author Contributions**: EJB, CL, MB, TC, MK, JA, DF, BKN participated in conceptualization. EJB, MK assisted in procuring the data and writing the first draft. EJB, MK generated the coding for the analyses. All authors reviewed and commented on subsequent drafts of the manuscript. **Funding**: NIH NIMHD K23MD017253.

## Abstract

**Background:** Naturally occurring dietary patterns, a major contributor to health, are not well described among those with cardiovascular disease (CVD) – particularly in light of socioeconomic vulnerability. We sought to identify major dietary patterns in the US and their distribution by CVD, social risk factors, and Supplemental Nutrition Assistance Program (SNAP) participation.

**Methods:** This was a cross-sectional study among 32,498 noninstitutionalized adults from the National Health and Nutrition Examination Survey (2009-2020). We used principal component analysis to identify common dietary patterns. Individuals were assigned to the pattern for which they had the highest component score. Using multinomial logit regression, we estimated the percentage whose diets aligned with each pattern in population subgroups stratified by CVD, social risk factors, and SNAP. Analyses were adjusted for age, gender, race and ethnicity, total energy intake, and year, with sampling weights to provide nationally representative estimates.

**Results:** Four dietary patterns were identified among US adults: American (33.7%; high in solid fats, added sugars, and refined grains), Prudent (22.6%; high in vegetables, nuts/seeds, oils, seafood, and poultry), Legume (15.8%), and Fruit/Whole Grain/Dairy (27.9%), that together explained 29.2% of dietary variance. More adults with prevalent CVD (37.1%) than without (33.3%, p=0.005) aligned with the American Pattern, with no differences among other patterns. Each additional social risk factor associated with more adults aligned with American (2.5% absolute increase) and Legume (1.3%), and fewer aligned with Prudent (-1.9%) and Fruit/Whole Grain/Dairy (-1.9%) patterns (p<0.001 each). Analysis of dietary patterns across SNAP participation showed higher proportion of SNAP participants and income-eligible SNAP non-participants compared to non-eligible adults for the American (40.2% [38.1, 42.3%], 35.1% [32.7, 37.5%], 31.9% [31.0, 32.8%], respectively) and Legume patterns (17.2% [15.6, 18.8%], 17.8% [16.1, 19.5%]), 15.4% [14.6,16.1%], respectively) and less for Prudent (17.0% [15.5, 18.6%], 20.2% [18.2, 22.3%], 24.2% [23.3, 25.1%], respectively) and Fruit/Whole Grain/Dairy Patterns (25.6% [23.8%, 27.3%], 26.9%[27.6%,29.5%], 28.6% [27.6%, 29.5%], respectively).

**Conclusions:** Empirical dietary patterns vary by CVD and socioeconomic vulnerability. Initiatives to improve nutrition in at-risk individuals should consider these naturally occurring dietary patterns and their variation in key subgroups.

## Introduction

Poor diet is among the greatest contributing factors to premature death and morbidity in the US, most commonly from cardiovascular disease (CVD).^1,2^ Socioeconomically vulnerable populations with food insecurity or other social risk factors also have higher burdens of chronic diseases such as CVD, at least partly related to differences in diet.^3–6^ These sociodemographic disparities in nutrition quality and CVD have also failed to improve over time, despite increased scientific understanding of diets and health and investments in government nutrition programs such as the Supplemental Nutrition Assistance Program (SNAP), a federal program with an annual budget of $100 billion.^7–11^ A better understanding of how dietary patterns vary by cardiovascular health and socioeconomic vulnerability may inform more effective clinical and public health interventions.

The nutritional value of diets can be evaluated by individual food groups, nutrients, or prespecified institutionally-derived dietary pattern scores (i.e. metrics derived to quantify adherence to a particular dietary pattern such as the Healthy Eating Index (HEI) that measures adherence to the US dietary guidelines, Mediterranean diet score, and DASH diet score).^10,12–17^ These institutionally-derived dietary scores were developed for generally healthy populations, rather than for individuals with prevalent disease or socioeconomic challenges. They also focus on idealized food consumption, rather than naturally observed patterns of food consumption. The assessment of empirically derived dietary patterns (combinations of foods that comprise an individual’s diet) provides a complementary paradigm to inform clinical and public health efforts, especially to improve health equity across groups at risk for adverse outcomes.^18–21^ However, few studies have assessed how prevalent disease or socioeconomic vulnerability are associated with observed empirical dietary patterns. This is an important gap because, due to structural barriers and cultural preferences, individuals with CVD or socioeconomic vulnerability may have dietary habits that are overlooked by applying institutionally-derived dietary patterns.

To address these important knowledge gaps, we characterized empirical dietary patterns among US adults using updated nationally representative data from the National Health and Nutrition Examination Survey (NHANES). We hypothesized that adherence to these dietary patterns would vary by CVD status, social risk factors, and SNAP participation.

## Materials and Methods

### Design, Setting, and Population

NHANES is a continuous program released in two-year cycles that assesses the health and nutrition of US adults and children. NHANES data are sampled using a four-stage cluster design to represent the non-institutionalized civilian population. We pooled data from 2009-2020 NHANES cycles to provide a large, contemporary sample (overall response rate = 61.3%). All participants provided informed written consent, completed personal and household questionnaires, and received a health examination in a Mobile Examination Center. The participants provided up to two days of interviewer-administered 24-hour dietary recall using the multiple-pass method. The first recall was obtained during the mobile exam and the second by telephone three to ten days later. Full details of sample design have been published.^22–25^ This current study using deidentified data was deemed exempt by the University of Michigan Institutional Review Board. This study followed the Strengthening the Reporting of Observational Studies in Epidemiology-Nutritional Epidemiology reporting guideline.^26^

We included all participants age ≥20 years (n=32,498). Individuals <20 years were excluded due to differences in determinants of dietary habits and the low prevalence of CVD compared with adults.

### Dietary Patterns

Relevant food categories (n=29) were identified from USDA Food Pattern categories and converted to standardized servings using the Food Patterns Equivalents Database, including cup equivalents (fruit, vegetables, dairy), ounce equivalents (grains, protein foods), teaspoon equivalents (added sugars), gram equivalents (solid fats and oils), and number of drinks (alcohol).^27^ To derive observed dietary patterns, we analyzed food categories using principal components analysis (PCA), a dimensionality reduction method that identifies patterns of variation across multiple variables (**Table 1**).^28^ Eigenvalues were calculated and plotted (**Supplementary Figure 1**), which flattened after the fourth dietary pattern. We focused on these four patterns. For each participant we calculated the HEI-2020 score (range 0-100), with higher scores reflecting more compliance with the Dietary Guidelines for Americans.^29^

**Table 1:**
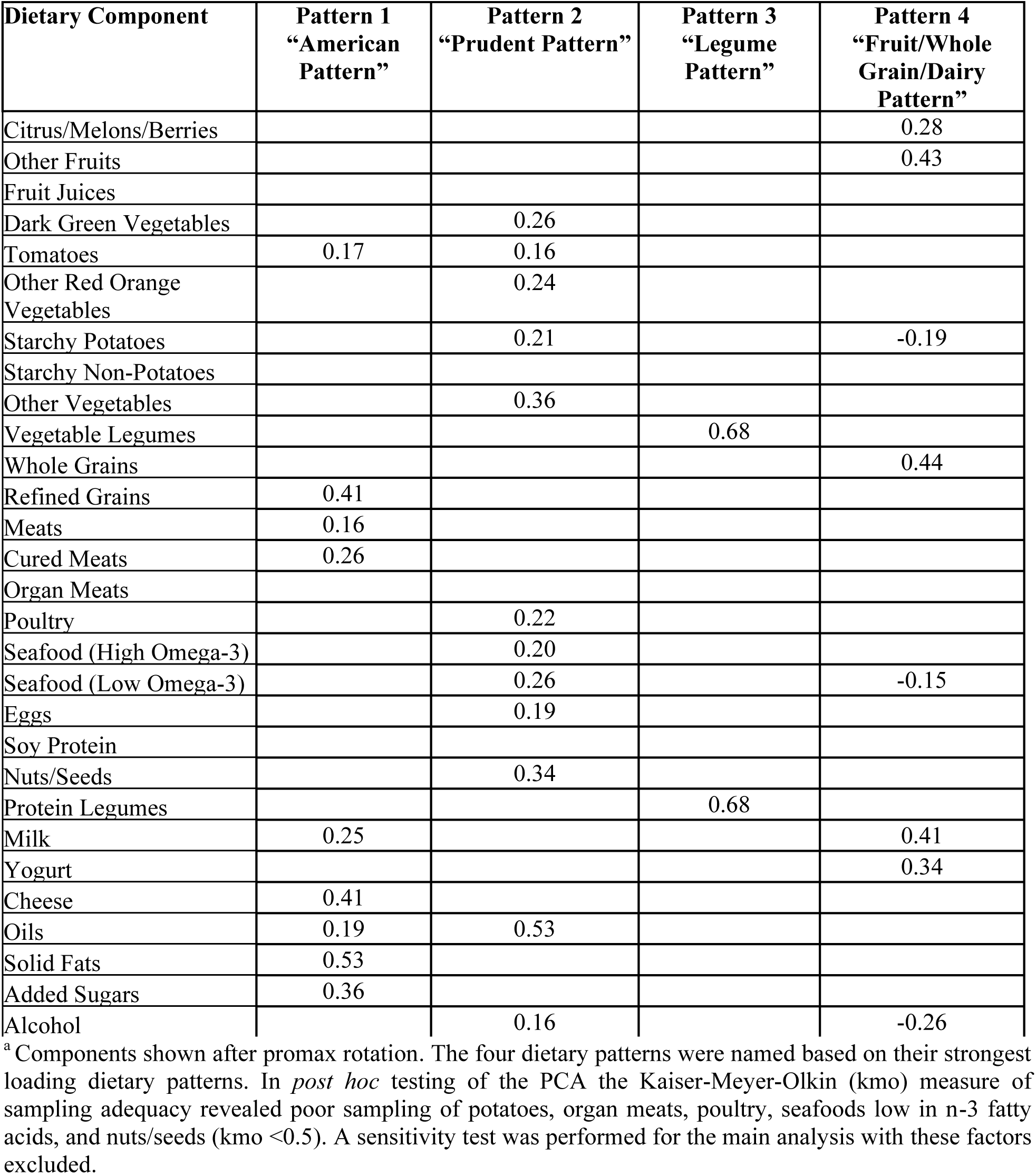
Principal Components Analysis Result Showing Component Loading (>0.15) of the Four Dietary Patterns ^a^.

| Dietary Component | Pattern 1<br>“American<br>Pattern” | Pattern 2<br>“Prudent Pattern” | Pattern 3<br>“Legume<br>Pattern” | Pattern 4<br>“Fruit/Whole<br>Grain/Dairy<br>Pattern” |
| --- | --- | --- | --- | --- |
| Citrus/Melons/Berries |  |  |  | 0.28 |
| Other Fruits |  |  |  | 0.43 |
| Fruit Juices |  |  |  |  |
| Dark Green Vegetables |  | 0.26 |  |  |
| Tomatoes | 0.17 | 0.16 |  |  |
| Other Red Orange<br>Vegetables |  | 0.24 |  |  |
| Starchy Potatoes |  | 0.21 |  | -0.19 |
| Starchy Non-Potatoes |  |  |  |  |
| Other Vegetables |  | 0.36 |  |  |
| Vegetable Legumes |  |  | 0.68 |  |
| Whole Grains |  |  |  | 0.44 |
| Refined Grains | 0.41 |  |  |  |
| Meats | 0.16 |  |  |  |
| Cured Meats | 0.26 |  |  |  |
| Organ Meats |  |  |  |  |
| Poultry |  | 0.22 |  |  |
| Seafood (High Omega-3) |  | 0.20 |  |  |
| Seafood (Low Omega-3) |  | 0.26 |  | -0.15 |
| Eggs |  | 0.19 |  |  |
| Soy Protein |  |  |  |  |
| Nuts/Seeds |  | 0.34 |  |  |
| Protein Legumes |  |  | 0.68 |  |
| Milk | 0.25 |  |  | 0.41 |
| Yogurt |  |  |  | 0.34 |
| Cheese | 0.41 |  |  |  |
| Oils | 0.19 | 0.53 |  |  |
| Solid Fats | 0.53 |  |  |  |
| Added Sugars | 0.36 |  |  |  |
| Alcohol |  | 0.16 |  | -0.26 |

<sup>a</sup> Components shown after promax rotation. The four dietary patterns were named based on their strongest loading dietary patterns. In *post hoc* testing of the PCA the Kaiser-Meyer-Olkin (kmo) measure of sampling adequacy revealed poor sampling of potatoes, organ meats, poultry, seafoods low in n-3 fatty acids, and nuts/seeds (kmo <0.5). A sensitivity test was performed for the main analysis with these factors excluded.

### Cardiovascular Disease and Socioeconomic Vulnerability

Prevalent CVD was defined as self-reported prior diagnosis of coronary artery disease, stroke, or heart failure from a health professional. Similar to a prior study,^30^ a social risk factor score (hereafter, social determinants of health (SDOH) score) was calculated based on having education less than high school graduation, being unmarried or not living with a partner, having Medicaid, other non-Medicare government health insurance, or being uninsured, experiencing food insecurity (low/very low food security), being unemployed (excluding retired individuals or students), or not having a routine location for healthcare. Unlike the prior study, we excluded income because SNAP participation is a similar marker. Housing was excluded due to systemic missingness from 2017-2020. One point was given for each factor and summed to calculate an overall score, with a higher score representing greater socioeconomic vulnerability. SNAP participation was defined as self-report of anyone in the household participating within the last 12 months. Income-eligible SNAP non-participants were defined by annual family income below 130% of the federal poverty level (FPL) (income-to-poverty ratio <1.3) without household SNAP participation within 12 months. SNAP non-eligible adults were those with annual family income ≥130% of FPL and no household SNAP participation within 12 months.

### Covariates

Demographic covariates included age, self-reported gender evaluated as a biologic variable, total energy intake (kcal/day), and race and ethnicity (Hispanic, non-Hispanic Asian (hereafter Asian, available from 2011 forward), non-Hispanic Black (hereafter Black), non-Hispanic White (hereafter White), and other). Race and ethnicity are sociocultural constructs that reflect a history of structural racism and cultural differences that were self-reported based on NHANES-defined categories.^22–25^

### Statistical Design and Analysis

Population characteristics overall and by CVD, SNAP status, and SDOH score were reported as percent (95% CI) for categorical variables and mean (SD) for continuous variables. Each individual was assigned to the dietary pattern for which they scored the highest. A single multinomial logit model examined the associations of four dietary patterns as the outcome with CVD status, SDOH scores, and SNAP status each evaluated as categorical variables. Marginal outputs from this model estimated the proportion of individuals aligned with each dietary pattern in each subgroup. All models were adjusted for age, gender, race and ethnicity, total energy intake, and NHANES survey cycle. Missing variables in NHANES (**Supplementary Table 1**) were multiply imputed, with results and analyses pooled from 20 imputed data sets.

We evaluated results overall and stratified by race and ethnicity. Statistical significance of interactions by race and ethnicity were tested by adding multiplicative interaction terms by CVD status, SDOH score, and SNAP status. A sensitivity analysis was conducted excluding food categories with Kaiser-Meyer-Olkin measure of sampling adequacy <0.5. All analyses utilized NHANES Mobile Examination Center weights to account for the complex survey design to produce nationally representative estimates. Adjustments for multiple comparisons were not performed because we were testing multiple separate hypotheses (see Introduction). Statistical significance was set at two-sided alpha=0.05. Analyses were performed using Stata v16 (StataCorp, LLC, College Station, TX).

## Results

### Population Characteristics

The study cohort included 32,498 adults representing 231 million US adults age ≥20 years (**Table 2**). Self-reported race and ethnicity included Asian (4.7%), Black (11.4%), Hispanic (14.9%), and White (65.0%). Prevalent CVD was present in 8.3%. Social risk factors were common: 14.8% had less than high school education, 37.5% not married or living with a partner, 34.6% publicly insured or uninsured, 15.7% with low or very low food security, 11.6% unemployed, and 15.6% lacking a routine location for healthcare. There were 17.0% SNAP participants, 10.2% income-eligible SNAP non-participants, and 72.8% non-eligible for SNAP.

**Table 2:** Weighted Population Characteristics (n=32,498, representing 230,612,714 US adults)

| Characteristic | %(95% CI) or mean (SD) |
| --- | --- |
| Age, years | 47.7 (17.1) |
| Female | 51.9% (51.3%,52.6%) |
| Race | - |
| Asian <sup>a</sup> | 4.7% (4.0%,5.6%) |
| Black | 11.4% (10.0%,13.1%) |
| Hispanic | 14.9% (13.0%,17.1%) |
| White | 65.0% (62.0%,67.8%) |
| Other <sup>a</sup> | 4.0% (3.5%,4.5%) |
| Cardiovascular Disease Diagnosis | 8.3% (7.8%,8.9%) |
| Social Determinants of Health | - |
| Not High School Graduate or GED recipient | 14.8% (13.7%,16.0%) |
| Not Married or Living with Partner | 37.5% (36.2%,38.9%) |
| Public Insurance or Uninsured | 34.6% (33.2%,36.1%) |
| Low, or Very Low Food Security <sup>b</sup> | 15.7% (14.8%,16.6%) |
| Unemployed | 11.6% (10.9%,12.4%) |
| No Regular Healthcare Access | 15.6% (14.8%,16.5%) |
| Adverse SDOH score | - |
| 0 | 34.5% (32.9%,36.1%) |
| 1 | 29.1% (28.3%,30.0%) |
| 2 | 18.7% (18.0%,19.5%) |
| 3 | 11.3% (10.6%,12.1%) |
| ≥4 | 6.4% (5.9%, 6.9%) |
| SNAP Participation Status | - |
| Non-eligible | 72.8% (71.1%,74.4%) |
| Income-Eligible Non-Participant | 10.2% (9.6%,10.9%) |
| Participant | 17.0% (15.7%,18.4%) |
| Total energy intake (kcal/day) | 3935 (1679) |

<sup>a</sup> Other included non-Hispanic Asian individuals in the 2009-2010 cohort.
<sup>b</sup> Measured using the US Household Food Security Survey Module.

In unadjusted analyses, those with a CVD diagnosis were more likely to be older, male, Black or White, less educated, unmarried or not living with a partner, food insecure, unemployed, have regular healthcare access, have a higher SDOH score, and be SNAP participants or income-eligible SNAP non-participants (**Supplementary Table 2**), highlighting the intersections of CVD with socioeconomic vulnerability. Those with a higher vs. lower SDOH score were younger and more likely to be male, Black, or Hispanic (**Supplementary Table 4**).

SNAP participants were younger, higher proportion female and Black individuals, and had a higher SDOH score (**Supplementary Table 3**), compared to non-eligible adults.

### Dietary Patterns

Among the four dietary patterns identified (see **Visual Abstract** and **Table 1)**, the “American” Pattern included higher intake of refined grains, meats, cured meats, milk, cheese, oils, solid fats, and added sugars (variance explained: 10.2%). The “Prudent” Pattern was defined by higher vegetables, meat, poultry, seafood, eggs, and oils, and lower milk (variance explained: 8.0%). The “Legume” Pattern had higher vegetables and legumes (variance explained = 6.5%). The “Fruit/Whole Grain/Dairy” Pattern included higher fruits, whole grains, soy protein, nuts/seeds, milk, and yogurt, and lower potatoes and eggs (variance explained = 4.6%). About one-third (33.7%) of US adults’ diets aligned most closely with the American, followed by Fruit/Whole Grain/Dairy (27.9%), Prudent (22.6%), and Legume (15.8%). Among individuals aligning with each pattern, the mean (SD) HEI score was highest for the Fruit/Whole Grain/Dairy pattern (62.0 [12.0]), followed by Prudent (58.2 [10.8]) and Legume (55.0 [12.5]) patterns, and lowest for the American pattern (42.4 [8.9]) (p<0.001).

### Dietary Patterns by Prevalent CVD

In the fully adjusted model, the proportion of US adults consuming diets aligned with the American pattern was higher among those with CVD (37.1% [95% CI: 34.6, 39.7%]) compared to those without (33.3% [32.4, 34.2%], p=0.005) (**Figure 1, Supplementary Table 5**). The proportion of adults aligned with each the other dietary patterns was similar among those with or without CVD.

**Figure 1:**
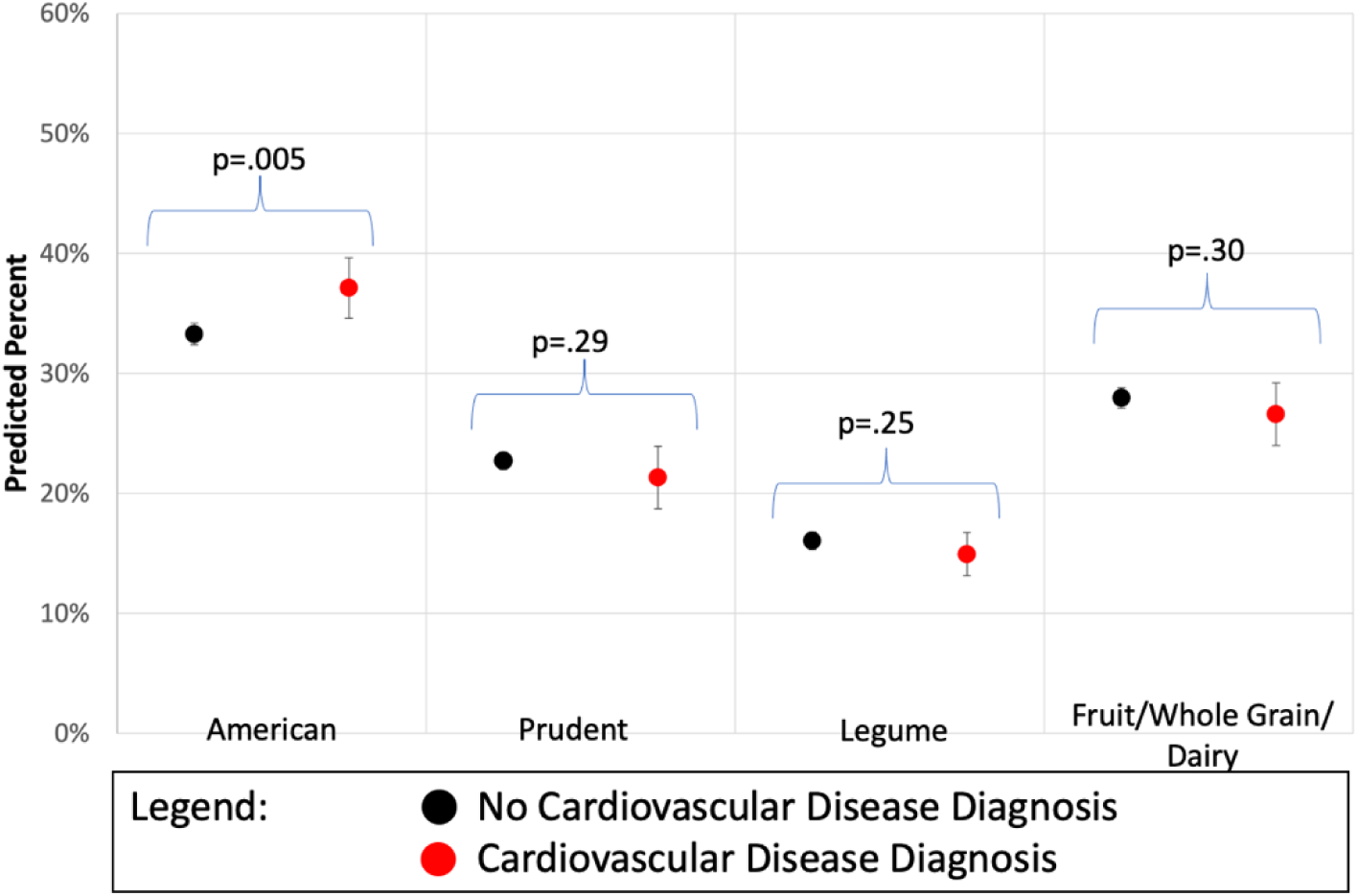
Probability of US Adults Following Four Different Dietary Patterns Across Cardiovascular Disease Status. Note: Proportions are predicted as part of a multinomial logit model.

### Dietary Patterns by SDOH score

The SDOH score was associated with all four dietary patterns. Those with more social risk factors were more likely to align with the American and Legume patterns, and less likely to align with the Prudent and Fruit/Whole Grain/Dairy patterns (p-trend<.001 for each, **Figure 2**, **Supplementary Table 6**). Each additional social risk factor was associated with a 2.5% absolute higher percentage of consuming the American, 1.3% higher consuming the Legume, 1.9% lower consuming the Prudent, and 1.9% lower consuming the Fruit/Whole Grain/Dairy Pattern.

**Figure 2:**
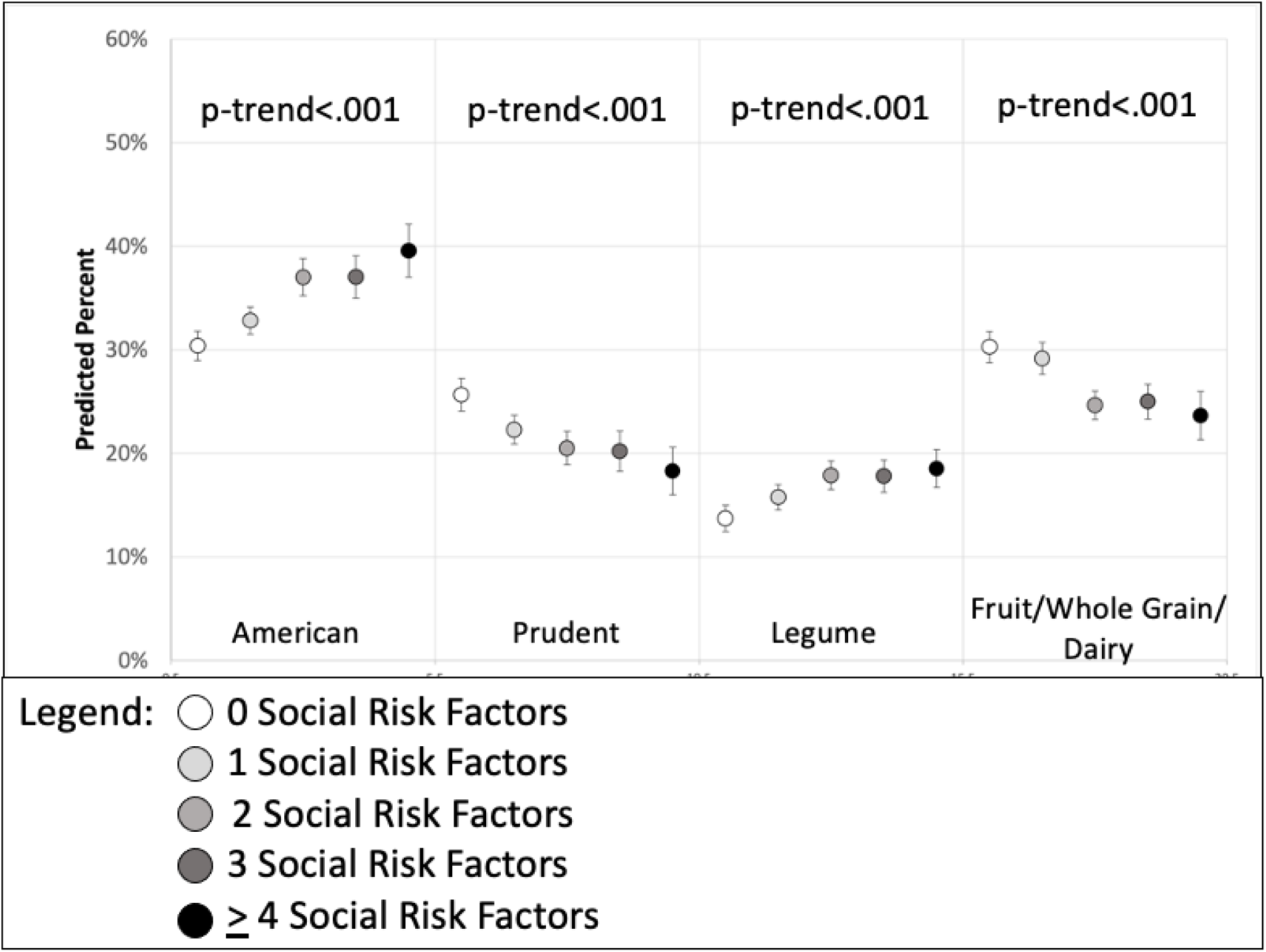
Probability of US Adults Following Four Different Dietary Patterns Scross Number of Social Risk Factors. Note: Proportions are predicted as part of a multinomial logit model.

### Dietary Patterns by SNAP Status

SNAP status was associated with all four dietary patterns. The proportion of US adults aligned with the American Pattern was higher among SNAP participants (40.2% [38.1, 42.3%]) than income-eligible SNAP non-participants (35.1% [32.7, 37.5%], p<0.001), and both were higher than non-eligible adults (31.9% [31.0, 32.8%], p<0.001 and p=0.01, respectively (**Figure 3, Supplementary Table 7**). In contrast, the proportion aligned with the Prudent Pattern was lower among SNAP participants (17.0% [15.5, 18.6%]) than income-eligible SNAP non-participants (20.2% [18.2, 22.3%],p=0.006), with both lower than non-eligible adults (24.2% [23.3, 25.1%], p<0.001, and p=0.001, respectively). The proportion aligned with the Legume Pattern was similar between SNAP participants (17.2% [15.6, 18.8%]) and income-eligible SNAP non-participants (17.8% [16.1, 19.5%]), and both higher than non-eligible adults (15.4% [14.6,16.1%], p=0.03, and p=0.009, respectively). The proportion aligned with the Fruit/Whole Grain/Dairy Pattern was lower among SNAP participants (25.6% [23.8%, 27.3%]) than non-eligible adults (28.6% [27.6%, 29.5%], p=0.003), and both were similar to non-eligible adults (26.9%[27.6%,29.5%]).

**Figure 3:**
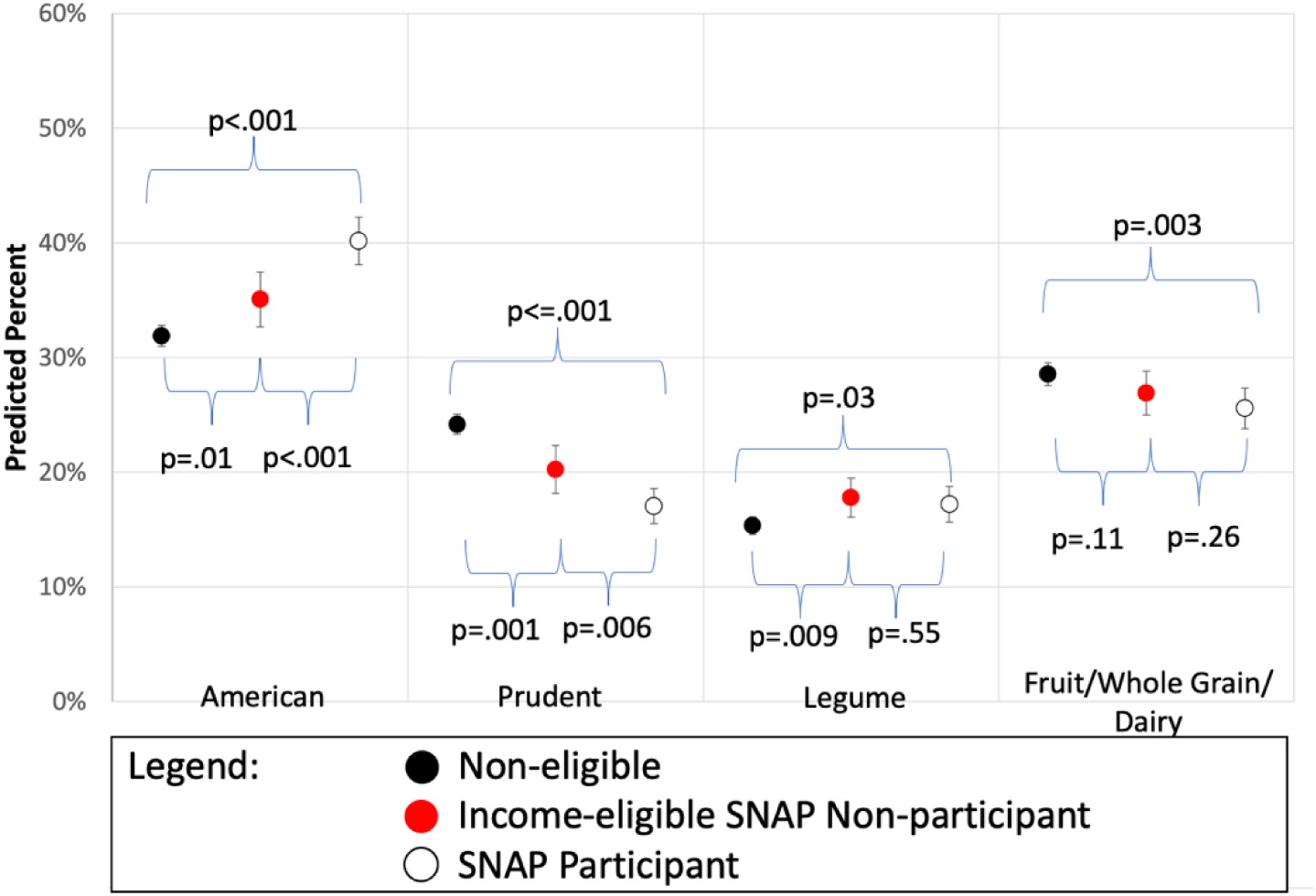
Probability of US Adults Following Four Different Dietary Patterns Across SNAP Participation Status. Note: Proportions are predicted as part of a multinomial logit model.

### Differences by Race and Ethnicity

The associations of prevalent CVD with dietary pattern were not significantly different by race and ethnicity (p-interaction=0.42; **Supplementary Table 5, Supplementary Figure 2**). The associations of SDOH score with dietary pattern was significantly different by race and ethnicity (p-interaction=0.002; **Supplementary Table 6, Supplementary Figure 3**). A key differences from the entire sample included a lack of association between increasing SDOH score and the American Pattern among Hispanic individuals. The associations of SNAP status with dietary pattern was significantly different by race and ethnicity (p-interaction=0.02; **Supplementary Table 7, Supplementary Figure 4**). Key differences from the entire sample included that SNAP status was not associated with differences in alignment to the Legume diet among White individuals or alignment with the American diet among both Hispanic and Asian individuals.

For coefficients of all covariates see **Supplementary Table 8**.

### Sensitivity Analyses

Findings were generally similar in sensitivity analyses excluding food categories not meeting measures of sampling adequacy (**Supplementary Tables 9 and 10**).

## Discussion

In this nationally representative study of US adults, we identified four predominant dietary patterns and differences in the distribution of these dietary patterns according to prevalent CVD and socioeconomic vulnerability. The least healthy American dietary pattern (HEI-2020 score 42.4) was most common overall and more common among those with prevalent CVD, higher SDOH score, or SNAP participating/income-eligible. Higher SDOH score and SNAP participating/income-eligible, but not prevalent CVD, was also associated with the other three empirically derived diet patterns. Adults having more SDOH or SNAP participating/income-eligible were less likely to follow the healthier Prudent dietary pattern (HEI-2020 score 58.2).

The healthier Fruit/Whole Grain/Dairy pattern (highest HEI-2020 score 62.0) was also less prevalent among those with higher SDOH score and SNAP participants. Interestingly, the Legume pattern (HEI-2020 score 55.0) was least common overall but *more* common among those with higher SDOH scores, SNAP participating, or SNAP income-eligible.

These new findings provide further support for the role of unhealthy diet as a contributor to higher risk for recurrent or incident cardiometabolic events in the setting of established CVD or socioeconomic vulnerability. From a health equity lens, this emphasizes that the most vulnerable individuals, from either a health or socioeconomic standpoint, may require more concentrated nutritional supports to prevent diet-related adverse outcomes. In contrast, socioeconomically vulnerable adults, in particular from Hispanic backgrounds, were more likely to align to a healthier dietary pattern marked by higher legume intake. Legumes have lower cost per serving than other protein foods, are rich in fiber and phytonutrients, and are associated with lower risk of CVD.^31–33^ This suggests that, in some cases, there may be healthier dietary patterns that are accessible among socioeconomically vulnerable individuals, which may be further shaped by social and cultural influences. These findings support the need for further investigation of the role of the Legume Pattern, especially for socioeconomically vulnerable individuals.

Our findings are also relevant for the burgeoning implementation of “Food is Medicine” approaches to integrate food-based nutritional therapies into healthcare.^34–36^ Understanding naturally occurring patterns of dietary intake by CVD and socioeconomic vulnerability can inform clinical interventions and practices to improve nutrition quality, health outcomes, and health equity for all Americans. This is especially important for groups at higher risk related to food insecurity, such as SNAP participants.^12,37–40^ Randomized trials have shown that healthier dietary patterns can reduce CVD events by 30-40%.^41–43^ Understanding how to introduce healthier food items within existent dietary patterns may facilitate sociocultural relevance of Food is Medicine programs, such as medically tailored groceries or meals; and better ways to incentivize healthier food selection and nutrition education within SNAP. Further, people with diet-sensitive chronic conditions may benefit from dedicated dietary counseling. This is proposed in the federal Medical Nutrition Therapy Act of 2023, which would expand Medicare coverage for dietary counseling to those with CVD and cardiovascular risk factors.^44–48^ This is sorely needed because dietary counseling is underutilized in the setting of CVD.^49^

Our investigation considered race and ethnicity as a sociocultural construct,^50^ whereby individuals may be more likely to follow dietary patterns due to cultural and regional influences and family heritages. Most of the findings in our study were similar across race and ethnicity.

Our findings are consistent with prior studies suggesting that a meaningful proportion of the relationships between socioeconomic factors, race and ethnicity, and life expectancy are mediated by behavioral and metabolic risk factors, including diet.^51^ Additional research into how dietary patterns are achieved and impacted in the context of socioeconomic vulnerability are needed to help us understand our few observed differences across race and ethnicity.

Our findings concur with prior findings in the context of CVD and socioeconomic vulnerability. In the prospectively collected Multi-Ethnic Study of Atherosclerosis cohort four empirical patterns were identified.^18^ In their study, a Fats and Processed Meat pattern (analogous to the American pattern in our study) was associated with higher CVD risk (HR of quintile 5 vs 1: 1.82 (95% CI: 0.99, 3.35). However, unlike the null finding in our cross-sectional study, a Whole Grain/Fruit pattern was associated with lower risk (HR 0.54 [0.33, 0.91]).^18^ In the Nurses’ Health Study two dietary patterns were found, with lower risk from a Prudent (HR 0.72 (0.60- 0.87)) compared to a Western/Processed food pattern.^19^ Studies considering participants in SNAP have found that participants consume fewer healthy foods and nutrients and have lower scores on institutionally-derived diet patterns, at least partly related to structural barriers faced in accessing and affording a healthful diet.^12,37,39,40,52,53^

Few prior studies have evaluated naturally occurring dietary patterns in nationally representative US populations. Among US cancer survivors, two dietary patterns were identified, including a Prudent pattern that was associated with lower food insecurity.^54^ Two studies identified patterns in nutrient intake in relation to hypertension and bone health, but did not evaluate food groups.^55,56^ One study from the early 1990s identified four dietary patterns among Mexican Americans in relation to gallbladder disease, including patterns marked by highly processed food, high vegetables, and high beans.^57^ Our study builds upon and greatly extends this prior work by identifying contemporary food patterns and their relations to CVD and socioeconomic vulnerability.

### Strengths

Study strengths include the large nationally representative cohort and racial and ethnic diversity, which support the reliability and generalizability of our findings. Sensitivity analyses to address shortcomings of the data did not change the results. Lastly, we used rigorous and established methods for the PCA that helps account for correlation between food categories.^58^

### Limitations

Potential limitations include the cross-sectional nature of the data source, which limits causal inference. Also, NHANES relies on individual self-reports of CVD, SNAP, and social risk factors, which may increase misclassification of these exposures. Dietary habits were assessed using the average of two 24-hour recalls, and longer dietary intake monitoring may increase the accuracy of observations.

## Conclusion

Empirical dietary patterns vary by both cardiovascular health and especially socioeconomic vulnerability. Initiatives to improve nutrition in at-risk individuals should consider these naturally occurring dietary patterns and their variation in key subgroups. Interventions in health policy and clinical practice should be evaluated to promote healthier dietary patterns among people with or at risk for CVD.

## Data Availability

Data are publicly available for download to the public. Coding used for the study are available upon request.

## Notes

**Conflicts of Interest/Disclosures**: EJB reports research funding from the National Institutes of Health (K23MD017253) and the Blue Cross Blue Shield of Michigan Foundation. He has received consulting fees from New Amsterdam Pharmaceuticals. Other authors report no conflicts of interest.

### Competing Interest Statement

EJB reports research funding from the National Institutes of Health (K23MD017253) and the Blue Cross Blue Shield of Michigan Foundation. He has received consulting fees from New Amsterdam Pharmaceuticals. Other authors report no conflicts of interest.

### Author Declarations

This current study using deidentified data was deemed exempt by the University of Michigan Institutional Review Board.

